# Evaluating the Quality of Mental Health Information Generated by Large Language Model Chatbots

**DOI:** 10.1101/2024.12.20.24319373

**Authors:** Gargi Porwal, Jitendra Jeenger

## Abstract

Artificial intelligence (AI) large language models (LLMs) hold great potential to transform psychiatry and mental health care by delivering relevant and tailored mental health information. This study aimed to evaluate the quality of mental health information generated by LLMs by determining their level of accessibility, reliability, and interpreting any bias present. Generative Pre-trained Transformer-4 (GPT-4) (San Francisco, California: Open AI), Gemini 1.5 Flash (Mountain View, California: Google LLC), and Large Language Model Meta AI 3.2 (Llama 3.2) (Menlo Park, California: Meta Inc.) were prompted with 20 questions commonly asked about anxiety and depression. The responses were evaluated using Flesch-Kincaid readability tests to quantify their ease of understanding through grade level and reading ease score measures. The text was subsequently analyzed using a modern DISCERN score to assess the reliability of the health-related information presented. Finally, the LLM responses were evaluated for stigmatizing language using communication and language guidelines for mental health. A significant difference in grade levels and reading ease scores was observed between GPT-4 and Llama (p < 0.01) and Gemini and Llama(p < 0.01), with both GPT-4 and Gemini having higher readability scores. No significant difference was observed in the grade level and reading ease scores between GPT-4 and Gemini (p > 0.05). All three models reported moderate reliability scores but no significant differences were observed (p > 0.05). GPT-4, Gemini, and Llama included stigmatizing phrases in 10%, 15%, and 20% of their responses respectively. These phrases were attributed to descriptions of mental health conditions and substance use; however, no significant differences were observed in the proportions of stigmatizing phrases present across the three models (p > 0.05). Furthermore, comparisons between anxiety and depression responses for each model revealed no significant differences in readability, reliability, or bias. All models demonstrated the ability to generate mental health information that generally satisfied criteria for accessibility, reliability, and minimal bias, with GPT-4 and Gemini reporting higher readability than Llama. However, the lack of additional resources cited in responses and the occasional presence of certain stigmatizing phrases, indicate that additional model training and fine-tuning, specifically for mental health applications, may be necessary before these tools can be deployed on a large scale for mental health care.

## Introduction

Globally, there is a mental health crisis, with half of all individuals expected to experience a mental health disorder in their lifetimes.^1^ Access to mental health care remains unable to meet this increasing demand due to several challenges, including a shortage of trained professionals, financial barriers, and stigma that prevents many individuals from seeking help.^2,3^ As societal gaps limit access to care, people are increasingly turning to digital tools to access mental health information and resources.^4^

Generative artificial intelligence (AI) large language models (LLMs) are quickly becoming one such tool. With their ability to interpret, summarize, and generate texts, LLMs have become popular tools for web-based information-seeking and have been utilized for mental health support by people experiencing distress but lacking access to immediate help. These tools provide support in various ways, reframing negative thoughts and emotions, reducing anxiety, guiding users through cognitive behavioral therapy (CBT) techniques, and providing crisis interventions.^5-7^

People often rely on online resources for quick and accessible health information, making LLMs valuable tools for scalable interventions that broaden access to mental health care. These AI models can be leveraged to deliver low-cost, always available, and anonymous advice and support. However, while LLMs hold promise for enhancing access to care and delivering personalized interventions, they also present risks, including generating hallucinations that may provide inaccurate or fabricated information, lacking predictability or interpretability, perpetuating inherent biases, and violating ethical concerns.^8^ Therefore, it becomes imperative to evaluate the clarity, accuracy, and safety of the mental health information generated by LLMs to ensure it promotes an accurate and effective understanding of mental health conditions and is understandable by diverse populations that may have varying literacy levels.

Given the opportunities and risks associated with LLMs, it is important to assess how models respond to common questions and concerns about mental health conditions, particularly anxiety and depression, which are among the most prevalent disorders.^9^ In this cross-sectional study, Generative Pre-Trained Transformer 4 (GPT-4), Gemini 1.5 Flash, and Large Language Model Meta AI (Llama) 3.2 were prompted with queries about anxiety and depression, and their responses were evaluated for readability, reliability, and presence of stigmatizing or biased language.

## Methods

Open AI’s GPT-4, Google’s Gemini 1.5 Flash, and Meta’s Llama 3.2 were each prompted with 20 common questions about anxiety and depression. The prompts were selected by the author (JJ), a practicing psychiatrist, based on an analysis of frequently asked questions sourced from online databases related to these mental health conditions.^10-14^ The questions focused on diagnosis and general information about anxiety and depression, treatment options, and guidance on accessing support resources for both conditions. Table 2 in Supplementary Materials provides a list of all the questions used in this study. For each question, a new chat window was initiated with prior history cleared, and the question was posed separately to each of the three LLMs. As LLMs may generate several answers for the same question in different instances, only the initial response provided for each question was recorded and examined.

The responses generated by the LLMs were analyzed using the Flesch-Kincaid calculator, which evaluated word count, sentence count, average words per sentence, and average syllables per word. These metrics were utilized to compute the Flesch-Kincaid grade level and reading ease scores, which quantify the readability through formulas that assign different weights to the total number of words, sentences, and syllables. Flesch-Kincaid reading ease ranges from 0-100, with higher scores indicating greater ease of understanding. The Flesh-Kincaid grade level similarly assesses the appropriate educational level to comprehend text, and a higher grade level suggests the content is more complex.

A modified DISCERN score assessed the reliability of each response.^15^ The tool utilized a five-question scale, with each question scored as 0 or 1 based on whether the response met specified criteria. A final score of 5 indicates high reliability, whereas a score of 0 suggests the text lacks scientific reliability. Three psychiatrists evaluated the responses generated by each LLM and graded their reliability using the modified DISCERN scale. The scores for each response were averaged and analyzed.

Responses were evaluated for biases using six principles derived from media mental health communication guidelines: stigma reduction, promotion of help-seeking behavior, avoidance of sensationalism, adherence to evidence-based content, person-centered language, and empowering language.^16,17^ Each response was rated as 0 or 1, where 0 indicated that the response contained at least 1 stigmatizing phrase, and 1 suggested that the response did not contain any stigmatizing language. The rating was performed by comparing each response against the list of preferred and problematic phrases listed in Mindframe’s guide, *Our words matter: Guidance of Language Usage*.^16^ Three psychiatrists rated the responses according to these guidelines, and the majority rating was used for further analysis.

### Statistical Analysis

A one-way ANOVA compared the mean Flesch-Kincaid grade level, reading ease score, and modified DISCERN score among GPT-4, Gemini, and Llama. Significant differences identified by the ANOVA test were further analyzed using a post-hoc Tukey HSD test for pairwise comparisons to determine which specific pairs among GPT-4, Gemini, and Llama showed statistically significant differences. An unpaired t-test compared the mean values of each variable for anxiety and depression responses within each model. A two-proportion Z-test was conducted to assess whether the adherence rates to communication guidelines significantly differed among the GPT-4, Gemini, and Llama models. Additionally, the test examined whether adherence varied between responses addressing anxiety and depression within each model. P-value < 0.05 was considered statistically significant.

## Results

### Text Readability

Significant differences were identified for mean word count, mean sentence count, and average syllables per word across responses generated by GPT-4, Gemini, and Llama, as shown in Table 1. Post-hoc pairwise comparisons revealed statistically significant differences in word count across all three models (GPT-4 and Gemini: p < 0.0001, GPT-4 and Llama: p < 0.0001, Gemini and Llama: p = 0.019). For sentence count, GPT-4 had a significantly higher mean compared to Gemini (p < 0.0001) and Llama (p < 0.0001), but no significant difference in sentence count was observed between Gemini and Llama. The average syllables per word were significantly higher for Llama than GPT-4 (p = 0.015), but no significant differences were found between GPT-4 and Gemini or Gemini and Llama. No significant differences were observed for the average number of words per sentence.

**Table 1:** Mean and Standard Deviation of Readability and Reliability Measures. * indicates p-value < 0.05 and the difference is considered statistically significant.

| Variables | GPT-4 | Gemini 1.5 Flash | Llama 3.2 | P-value |
| --- | --- | --- | --- | --- |
|  | Mean (SD) | Mean (SD) | Mean (SD) |  |
| Word Count | 424.6 (144.86) | 182.25 (65.69) | 266.25 (43.25) | $< 0.0001^*$ |
| Sentence Count | 46.95 (18.23) | 20.85 (9.71) | 25.65 (6.49) | $< 0.0001^*$ |
| Average Words per Sentence | 9.4 (1.96) | 9.38 (2.57) | 10.81 (2.35) | 0.0904 |
| Average Syllables per Word | 1.95 (0.15) | 1.98 (0.15) | 2.08 (0.13) | 0.01485* |
| Flesch-Kincaid Grade Level | 11.03 (1.93) | 11.3 (1.78) | 13.1 (1.95) | 0.001768* |
| Flesch-Kincaid Reading Ease | 32.75 (12.69) | 30.66 (12.19) | 20.32 (11.74) | 0.004531* |
| modified DISCERN | 3.2 (0.61) | 3.15 (0.81) | 2.95 (0.69) | 0.5412 |

There were significant differences in the Flesch-Kincaid grade level and reading ease scores (Table 1). Llama had a significantly higher grade level than both GPT-4 (p = 0.002) and Gemini (p = 0.011), and a lower reading ease score than GPT-4 (p = 0.006) and Gemini (p = 0.025) (Figure 1A, 1B). GPT-4 had a lower grade level and higher reading ease score than Gemini, but these differences were not statistically significant.

Comparisons between responses addressing anxiety and depression for each model revealed that Gemini generated significantly higher word (p = 0.005) and sentence counts (p = 0.048) for anxiety-related responses compared to those for depression. Similarly, GPT-4 produced significantly higher average words per sentence for anxiety responses than for depression (p = 0.025). For each LLM, the grade level was higher, and the reading ease was lower for anxiety responses than for depression responses, but these differences were not statistically significant. No significant differences were observed in average syllables per word for any of the models.

### Text Reliability

GPT-4 had a higher mean modified DISCERN score than Gemini and Llama, but these differences were not statistically significant (Table 1). Although modified DISCERN scores were higher for anxiety prompts compared to depression prompts for all three models, these differences were not significant (Figure 1C).

LLMs were able to effectively answer each prompt and cite reliable sources used to generate responses. The most common DISCERN criteria the LLMs failed to meet was addressing areas of uncertainty in their responses. 85% of the responses for each model did not meet this criterion. Listing additional sources of information for patient reference was another factor that the LLM tools failed to meet adequately. GPT-4 and Gemini cited additional references in 35% of the responses, while Llama did so for only 15%.

### Communication Guideline Adherence

Although the relevance of each guideline varied for different topics addressed in the prompts, some common patterns were observed in the responses. Responses employed person-centered and empowering language, and consistently included disclaimers emphasizing the importance of seeking professional help. Phrases such as “seeking help is a sign of strength, not a sign of weakness” and “many people with anxiety lead fulfilling lives” were observed across all three models, with particular emphasis on mental health conditions being treatable.

Despite these patterns, however, the LLMs were not able to incorporate non-stigmatizing language consistently throughout all their responses. Stigmatizing phrases appeared in 10% of GPT-4’s responses, 15% of Gemini’s responses, and 20% of Llama’s responses. This was especially evident in phrases describing anxiety and depression, with GPT-4 using the term “impairing”, and Gemini and Llama both describing the conditions as “debilitating.” Information about alcohol and substance use also included the terms “substance abuse” and “misuse.”

The differences in the proportion of responses with stigmatizing language were not statistically significant. Additionally, no significant differences were observed in the use of stigmatizing language in responses related to anxiety and depression across the models (Figure 1D).

**Figure 1:** Comparison of GPT-4, Gemini, and Llama in (a) Mean Grade Level, (b) Mean Reading Ease Score, (c) Mean Modified DISCERN Score, and (d) Proportion of Responses Adhering to Communication Guidelines for queries about anxiety and depression.

## Discussion

LLMs have the potential to transform population mental health by enhancing early diagnosis, personalizing treatments and interventions, and broadening access to care. Therefore, it is essential to assess the quality of information provided by these tools to determine whether it is accurate, unbiased, and understandable by diverse populations. This study evaluated the readability and reliability of information provided by GPT-4, Gemini, and Llama in response to common questions about anxiety and depression.

GPT-4 had a significantly higher word count and sentence count compared to both Gemini and Llama. This suggests the model may have been trained to provide longer and more detailed responses. However, this pattern did not result in GPT-4’s responses being more difficult to read or overly complex in structure or language. Responses generated by Llama had a significantly higher grade level and lower reading ease score than GPT-4 and Gemini, indicating that the readability of mental health text generated by Llama had significantly lower readability. Overall, GPT-4 and Gemini generated responses suitable for a high school audience, while Llama’s responses were more appropriate for a college-level audience. Considering that mental health information must be accessible to diverse populations, particularly underserved communities where health literacy rates are low, GPT-4 or Gemini may be better suited for mental health applications compared to Llama as they generate comprehensive responses with high readability. However, further training may be required to ensure the information is accessible by individuals from different backgrounds and literacy levels, especially since the average American is expected to be able to read at a 7^th^ or 8^th^ grade level.^18^ These findings were similar to Amin et al., which reported that LLMs were unable to generate health education material below a high school level without engineered prompting.^19^

The information generated by LLMs regarding the cause of anxiety and depression, the chronicity and recurrence of the conditions, and treatment options were verified with scientific journals and medical websites, indicating the information was generally accurate. However, when asked about common treatments for anxiety and depression, GPT-4 and Llama included alternate therapies such as acupuncture and herbal supplements among the treatment options. However, the existing literature presents mixed evidence regarding their efficacy, suggesting the material presented by these tools may not always be reliable, and consulting expert sources will be necessary.^20,21^ LLMs are trained on texts from a wide range of sources, including scientific journals, medical websites, and publicly available content, and lack the ability to discriminate the strength and quality of the data they are trained on. As a result, the information presented may include varying levels of scientific evidence, increasing risks of harm.^22^ This highlights a key challenge to ensure LLMs consistently provide clinically proven and empirical information in healthcare settings.

GPT-4 and Gemini provided relevant and up-to-date resources for anxiety and depression when explicitly prompted. However, Llama failed to incorporate the updated 988 phone number for the National Suicide Prevention Lifeline in its responses, instead referencing the previous number (1-800-273-TALK(8255))^23^, suggesting that the model may not have been trained on the most current mental health resources. Furthermore, Llama failed to provide direct links to online mental health resources, limiting the accessibility of the mental health support it offered. Consequently, GPT-4 and Gemini were considered more suitable for delivering information on mental health resources than Llama.

All three models often lacked additional resources or references in their responses when not explicitly prompted to cite them, which could be a critical limitation in the context of mental health. Individuals in distress benefit greatly from access to external resources they can turn to for further support. Providing such resources is crucial for making mental health education and care more accessible and effective. This suggests that simple queries, such as the ones used in this study, could not generate tailored responses and further model prompt engineering may be necessary to personalize responses.

The tone and style of the language in the generated responses were evaluated against communication guidelines for mental health. All models emphasized the importance of seeking professional help and incorporated person-centered, empathetic empowering language into their responses by consistently underscoring that anxiety and depression were treatable and that there were multiple avenues available for help. However, stigmatizing language was occasionally observed in descriptions of mental health conditions and substance use, indicating that societal biases and stereotypes may be inherent in the training data for LLMs. These findings corroborated previous studies that reported GPT-4 includes stigmatizing language when prompted to generate patient education materials on mental health and substance use.^24^ Further training to both identify and mitigate such biases will be necessary to ensure LLMs communicate mental health information in a more inclusive manner.

These findings highlight the opportunities presented by generative AI in the mental health domain. LLMs such as GPT-4 have demonstrated the ability to generate quality mental health information that is accessible, easily understandable, and person-centered. However, critical areas for refinement need to be addressed to ensure their outputs are consistently reliable and unbiased.

Drawbacks in the responses highlight the need to further improve LLMs for mental health applications. Fine-tuning models by providing extensive training on data-driven and research-based mental health information can improve LLM accuracy and reliability. Fine-tuning adjusts the LLM’s internal parameters, and training the model on scientific sources for mental health can refine the model’s network to home in on learning evidence-based content pertaining to mental health applications. Cultural humility and personalization can also be introduced through this mechanism by incorporating mental health information specific to different races, ethnicities, genders, and geographies in the training data. This can help ensure LLM responses are unbiased and tailored to the background of the user interacting with it. However, fine-tuning tasks can be computationally expensive and, therefore, may have limited scalability. Prompt engineering offers another avenue to improving LLM task performance that is accompanied by less computational costs. This technique may serve as an effective approach to improve model responses, especially since LLMs are sensitive to words or phrases used in prompts, often providing different responses to minor modifications in questions.^22^ This degree of unpredictability can pose risks in mental health contexts, but effective prompt engineering can train models to consistently provide reliable responses despite slight variations in queries. In fact, Maharjan et al. has shown that prompt engineering has been as effective as fine-tuning to improve LLM performance for medical question-answering.^25^ Through techniques such as in-context learning, LLMs can be trained on example queries and learn to generate context-appropriate responses with the desired style, tonality, content, and literacy level.^26,27^

### Limitations

The scope of this study posed some limitations that should be considered. Only 20 questions about anxiety and depression were used in this study. This limited set of prompts may not completely capture the variability in LLM performance on different topics and cannot be generalized to the broader range of mental health conditions. Additionally, the questions were selected from online datasets of common questions, which may not reflect the different ways individuals’ phrase or express their questions based on factors such as literacy levels and personal experiences. Similarly, evaluating the LLM responses using standardized measures of readability, reliability, and bias was valuable in quantifying these aspects of the text, but did not consider the variable understanding and reactions people may have in real-world settings depending on their educational and cultural backgrounds. The evaluation of bias was also constrained to language and common stigmatizing phrases without considering gender, race, ethnicity, and socioeconomic factors that might also impact the quality of responses. Future studies should focus on exploring these areas of bias in LLM-generated text and include the perspectives of individuals from diverse cultural and socioeconomic backgrounds when evaluating LLM responses. In addition to improving accessibility and accuracy, further evaluation of whether LLMs adhere to ethical standards and respect user privacy is crucial before their widespread adoption for mental health applications.

## Conclusion

This study evaluated the efficacy of LLMs (GPT-4, Gemini, and Llama) in addressing common questions about anxiety and depression and assessed both the readability and reliability of the responses, while also interpreting any bias present. Overall, the models generated quality responses that had varying levels of readability, with GPT-4 and Gemini presenting responses that were most readable and accessible. No significant differences in Flesch-Kincaid grade level or reading ease were observed between responses for anxiety and depression for any model, indicating that the models performed consistently across these two mental health conditions.

All three models provided information that was generally accurate; however, some of the responses, especially from Llama, had limited information about additional resources, raising concerns about the reliability of these tools for direct deployment in mental health applications. Additionally, some responses contained stigmatizing phrases about mental health, highlighting the need to improve the language of LLMs when responding to mental health concerns. Overall, LLMs offer significant opportunities to enhance the accessibility of mental health care, with GPT-4 and Gemini being more suitable for delivering relevant mental health information. But it will be critical to ensure they are further trained for mental health purposes, so they adhere to ethical standards and propagate accurate and reliable information.

## Data Availability

All data produced in the present study are available upon request to the authors

## Acknowledgments

We appreciate the contribution of three participating psychiatrists who independently evaluated the reliability and levels of bias of GPT-4, Gemini, and Llama’s responses to questions about anxiety and depression.

## Notes

### Competing Interest Statement

The authors have declared no competing interest.

### Funding Statement

This study did not receive any funding

### Summary of Updates

Updated Title, and changed authorship because one of the original authors didn't meet the criteria for authorship so they were acknowledged in the acknowledgements section instead.

