## Supplemental Table 2 for "Evaluating the Quality of Mental Health Information Generated by Large Language Model Chatbots"

**Questions**

What does anxiety feel like?

How can you tell if your anxiety is normal or a sign of anxiety disorder?

What is the difference between an anxiety attack and a panic attack?

When should someone seek treatment for anxiety?

What are common treatment options for anxiety?

Is medicine needed to treat anxiety?

Will someone struggle with anxiety their whole life?

What are helpful resources for anxiety?

How is anxiety diagnosed?

What causes anxiety disorders?

What does depression feel like?

What is the difference between depression and sadness?

What causes depression?

When should someone seek treatment for depression?

What are common treatment options for depression?

Is medicine needed to treat depression?

Will someone struggle with depression their whole life?

What are helpful resources for depression?

How is depression diagnosed?

How can you tell if someone is considering suicide and how to intervene?

**Table 2: Input Prompts to LLMs**
